# Comparing Narrative Storytelling Ability in Individuals with Autism Spectrum Disorder and Fetal Alcohol Spectrum Disorder

**DOI:** 10.1101/2022.09.20.22280005

**Authors:** Linh N. H. Pham, Adrian KC Lee, Annette Estes, Stephen Dager, Susan J. (Astley) Hemingway, John C. Thorne, Bonnie K. Lau

## Abstract

**Purpose:** Narrative discourse, or storytelling, is used in daily conversational interaction and reveals higher level language skills that may not be well captured by standardized assessments of language. Many individuals with autism spectrum disorder (ASD) and fetal alcohol spectrum disorder (FASD) show difficulty with pragmatic language skills and narrative production offers one method of assessing expressive and pragmatic language skills in an ecologically relevant manner. This study investigated narrative abilities on both local and global levels of adolescent/young adults with ASD and FASD, and their age- and sex-matched comparison group.

**Method:** Narratives from forty-five adolescents/young adults, 11 with ASD, 11 with FASD, 23 age- and sex-matched neurotypical comparison group, were elicited using a wordless storybook. They were then transcribed orthographically, formatted to the Systematic Analyses of Language Transcript (SALT) convention, and scored based on the narrative scoring scheme (NSS).

Additional analyses investigated local language measures such as the number of mental state and temporal relation terms produced, as well as global language measures including the mean length of utterance, total number of different words, total number of words, total number of utterances, rate of speech, and the narrative scoring scheme total score.

**Results:** On local language measures, no significant group differences were found. On global language measures, many aspects of narrative production in the ASD and FASD groups were comparable to each other and to the comparison group, although important differences were observed for the total number of words produced and rate of speech.

**Conclusions:** Given significant variability observed within groups, these findings suggest that language abilities should be assessed at an individual level. Future research should also consider additional variables that influence narrative production such as motivation, distractibility, or decision-making of individual subjects.

---

Narrative discourse is an important area of research in neurodevelopmental disabilities such as autism spectrum disorder (ASD; Barnes & Baron-Cohen, 2012; Colle et al., 2008; Losh & Gordon, 2014) and fetal alcohol spectrum disorder (FASD; Ganthous et al., 2017; Thorne et al., 2007) because the narrative skills required to tell a story include essential components of social communication and linguistic skills needed for daily conversational interactions. Due to the high variability in language skills and difficulty in assessing social communication, it is challenging to identify and characterize specific language impairments in individuals with ASD and FASD (Adams, 2002). While language impairment is not included in the current diagnostic criteria for ASD, deficits in social communication and social interaction, as well as restricted behaviors, interests, and activities, are indicated (American Psychiatric Association, 2013).

However, there is an increased incidence of language deficits in autistic individuals, which have been linked to learning and social difficulty (Tager-Flusberg & Sullivan, 1995). Similarly, language impairment is also not part of the diagnostic criteria for FASD (American Psychiatric Association, 2013), but is one of the more commonly reported associated impairments (Coggins et al., 2003; Thorne et al., 2007). Social communication is usually measured through two approaches: standardized assessment of language and parent/caregiver report. However, prior studies have identified limitations with these assessment methods such as difficulty in using standardized testing for children with ASD due to different reasons such as lack of motivation (Koegel et al., 1997) or the presence of bias in caregivers’ reports of child’s language (Tomasello & Mervis, 1994). Thus, narrative discourse offers an additional ecologically valid method of assessing real-world language skills and social communication in autistic individuals (e.g., Levinson et al., 2020) and FASD (e.g., Proven et al., 2014).

## Narrative Storytelling

Narrative discourse is a form of communication aimed at constructing a shared experience between the storyteller and listeners. Since it requires an exchange of information between speakers, it demonstrates speakers’ language and social communication skills, including the ability to receive, maintain, send, and comprehend verbal and non-verbal cues. For example, in narrative discourse, speakers need to maintain the topic of conversation and account for the perspective of the listener to provide informative utterances with the appropriate amount of detail.

Narrative analysis is a useful tool for revealing communication breakdown, qualitative differences in language, and information from dynamic social environments, which is often lost in the more formalized context of standardized language assessment. For example, the ability to tell a cohesive story with meaningful content or the ability to indicate the mental states of another person may indicate how well one can use language skills in real-world communication, beyond what is measured in standardized language assessments (Colle et al., 2008).

Prior studies have revealed that narrative discourse can provide additional information on language abilities in autistic individuals (Charman, 2004; King & Palikara, 2018; Manolitsi & Botting, 2011; Peristeri et al., 2017). For example, for some autistic children, standardized assessments required skills that did not adequately elicit the language skills relevant to their autistic diagnosis (Charman, 2004). In other words, the challenge with standardized assessment is the uneven profile of language competency across autistic children and the differences between language performance on measures of language competency. Although many autistic adolescents score within the average range on standardized assessments of language, higher-level language skill deficits may not be revealed by these assessments (King & Palikara, 2018). Similarly, one study found that autistic children showed receptive language deficits but not expressive language on standardized tests compared to children with specific language impairment; however, subsequent narrative assessment revealed deficits in expressive language involving story-telling and referencing (Manolitsi & Botting, 2011).

As children grow older, their narrative abilities also continue to develop. Specifically, they begin to produce rich, well-constructed narratives that include temporal and causal connections as well as evaluative references to each character’s mental states and emotions. As such, to tell a coherent narrative with a common theme throughout the story, children need to structure the story in an organized manner and use different devices such as temporal references to link together scenes, sentences, clauses, and prepositions (Karmiloff-Smith, 1985). The development of narrative competence, therefore, can be measured through self-generated narrative storytelling or story retelling, which have both been used to study the narrative skills of children and adults with different neurological conditions (storytelling; Colle et al., 2008; Losh & Gordon, 2014; Thorne et al., 2007; story retell, Diehl et al., 2006; Losh & Gordon, 2014; Novogrodsky, 2013).

One of the main differences between storytelling and story retelling is the complexity of the task that is required of the storytellers. While narrative story retelling only requires the storyteller to restate information from the previously heard or read story, in narrative storytelling, storytellers need to organize story events and structure them into a comprehensive narrative without the help of a model. Although narrative story retelling can be used to demonstrate one’s ability to identify relevant information and integrate the information into a coherent narrative, generating a narrative independently can be a more challenging (Kaderavek & Sulzby, 2000). One of the reasons is that the storytellers are expected to take in and constantly update the perspectives of the listeners while delivering the story, which can be achieved through referential expressions, connecting sentences and statements with transition words, and making reference to the mental states of the characters. Narrative storytelling, therefore, reflects the real-world interaction required in everyday educational and social settings.

## Narrative abilities in autism spectrum disorder

Although language impairment is not included in the current diagnostic criteria for ASD (American Psychiatric Association, 2013), in past studies of narrative production, autistic individuals were found to produce shorter and less complex sentences and stories (Banney et al., 2015; King et al., 2014; Tager-Flusberg & Sullivan, 1995), score lower on local language measures by producing fewer personal pronouns, temporal expressions, referential expressions (Colle et al., 2008; King et al., 2014; Novogrodsky, 2013), and causal statements (Siller et al., 2014; Tager-Flusberg & Sullivan, 1995). Autistic individuals produce stories that are biased toward local details (Barnes & Baron-Cohen, 2012). For example, an autistic individual may provide more specific details of the key component as opposed to the big picture that carries the critical elements of a story such as setting, character, and resolution. During a narrative task describing a film, participants with ASD were more likely to describe the setting by talking about the details in the setting such as describing the computer in the background of the scene instead of the overall setting such as a hospital or an office building as described by the control group. These global details are needed to capture the “big picture” of the story, which provides meaningful answers to each story element. Thus, autistic individuals may provide details about the setting or characters but may leave out significant information that helps a listener glean meaning from the information (Barnes & Baron-Cohen, 2012).

In contrast, when groups are matched on age and cognitive abilities, many studies report fewer differences between narratives produced by autistic individuals and neurotypical comparisons (Colle et al., 2008; Diehl et al., 2006; Losh & Capps, 2003; Losh & Gordon, 2014; Tager-Flusberg & Sullivan, 1995). While some past literature reported shorter and less complex narratives (Tager-Flusberg & Sullivan, 1995), other research demonstrates that the narratives produced by autistic individuals are comparable to comparison groups (Losh & Capps, 2003).

Pragmatic skills in highly verbal autistic individuals compared did not differ from age-, sex-, and IQ-matched comparison group; however, a narrative storytelling task revealed specific pragmatic deficits in autistic individuals such as using fewer personal pronouns, temporal expressions, and referential expressions (Colle et al., 2008). These findings suggest that narrative storytelling might be an important methodology to understand the challenges faced by autistic individuals.

## Narrative abilities in fetal alcohol spectrum disorders

Fetal alcohol spectrum disorder (FASD) results from prenatal alcohol exposure and is characterized by an array of physical, cognitive, social, and communication impairments. One of the common impairments in individuals with high levels of prenatal alcohol exposure that has been documented is difficulty in the use of language. As such, similarly to those with ASD, individuals with FASD also struggle with social communication and engaging in appropriate interpersonal communication (Coggins et al., 2003). Studies have shown speech production difficulties with both expressive and receptive language in some individuals with FASD (Aragón et al., 2008; McGee et al., 2009). Thorne et al. (2007) conducted a study analyzing stories for semantic elaboration and reference strategy for children ages 8-11 with FASD and found that the rate of ambiguous nominal reference was a marker to distinguish children with FASD from age-matched children without FASD. This finding indicated errors in referential cohesion, which refers to the failure to introduce and maintain references in an organized way that allows the listener to recognize references and follow through with the story. For example, a storyteller might decide to keep or reintroduce an object, such as “jar” for “bottle”, which is within the same semantic category of glass container. However, although the storyteller might reasonably expect the listener to recognize the overlapping nature of a *jar* and a *bottle*, in a decontextualized discourse, the listener might not be able to recognize. Additionally, consistent with the findings of Thorne et al. ‘s (2007), narrative discourse provides meaningful clinical tools to examine communication in individuals with FASD. In addition to referential cohesion, a study by Ganthous, Rossi, and Giachetti (2017) focused on fluency in oral narrative and found deficits on both global and local measures including structural elements of the storytelling as well as diverse vocabulary, length of sentence, and number of incomplete sentences.

With the limited number of studies available, there is a need for more research on narrative discourse in individuals with FASD. This is especially true given that prior studies have noted similar difficulties in social skills and communication for children with FASD and ASD (Bishop et al., 2007; Stevens et al., 2013), suggesting that narrative discourse may be a useful tool for evaluating individuals with FASD as it has proven to be for ASD. Moreover, while there are numerous studies demonstrating difficulty producing cohesive narratives in autistic children, very few have included adults (Beaumont & Newcombe, 2006; Colle et al., 2008; Geelhand et al., 2020). To better understand the role of narrative discourse in language ability, this study intends to investigate the narrative abilities of adolescent/young adults with ASD and FASD, as well as an age- and sex-matched comparison group. Therefore, the goal of the present study is to provide empirical research on the similarities and differences between ASD and FASD groups in terms of narrative skills and to build on prior research by quantifying the narrative abilities elicited for both local and global language measures. Whereas the global level of language demonstrates the speaker’s general understanding of the world and appropriate use of language form, the local level of language demonstrates the speaker’s ability to integrate and form cohesive narratives at the utterance level through linking within and across sentences. The local measures include enriched evaluative devices such as temporal references and mental states. The global measures include mean length of utterance, semantic measures of the number of used word roots, length of narrative (total number of words), total number of utterances, rate of speech, and scores of the Narrative Scoring Scheme (NSS) for each story (Miller et al., 2008). On the local language measures, while some past literature reported limited reference to internal states and temporal relations in autistic individuals (Baixauli et al., 2016; Colle et al., 2008), other studies did not find such differences (Losh & Capps, 2003; Tager-Flusberg & Sullivan, 1995). On the global language measures, while some past literature reported shorter lengths of stories (Tager-Flusberg & Sullivan, 1995) and worse narrative performance on story cohesiveness (Baixauli et al., 2016), others did not find such differences between ASD and neurotypical groups (Colle et al., 2008). Given the variability in findings, the second goal of the current study is to examine narrative storytelling as a tool to identify subtle language differences in the narrative ability of individuals with ASD and FASD that may not be captured by standardized testing. These analyses were intended to further clarify the key constructs that each measure contributed to in oral communication, particularly, storytelling.

This study addresses three specific questions: First, are there differences in the local and global structure of narratives produced by participants with ASD, FASD, and the comparison group? We hypothesized that participants with ASD and FASD would perform worse on the local measures but show no differences in the global measures in comparison to the neurotypical group. We further hypothesized that the FASD group would score higher on the local measures than the ASD group. Second, do the three groups differ in their use of terms that refer to temporal relations? We hypothesized that participants in the ASD and FASD groups would produce fewer temporal relation terms compared to the comparison group and the ASD group would produce the least of the three groups. Third, do the three groups differ concerning their use of terms that refer to mental states? We hypothesized that individuals with ASD would demonstrate the most difficulty in producing terms that referred to mental states compared to the FASD group and the comparison group.

## Methods

### Participants

Forty-five subjects participated in the study from four groups: individuals with ASD (*n* = 11), individuals with FASD (*n* = 11), and two comparison groups of neurotypical individuals (*n* = 23) that were age and sex matched to ASD and FASD groups and were later combined into one comparison group (see Table 1 for demographic information).

**Table 1.**
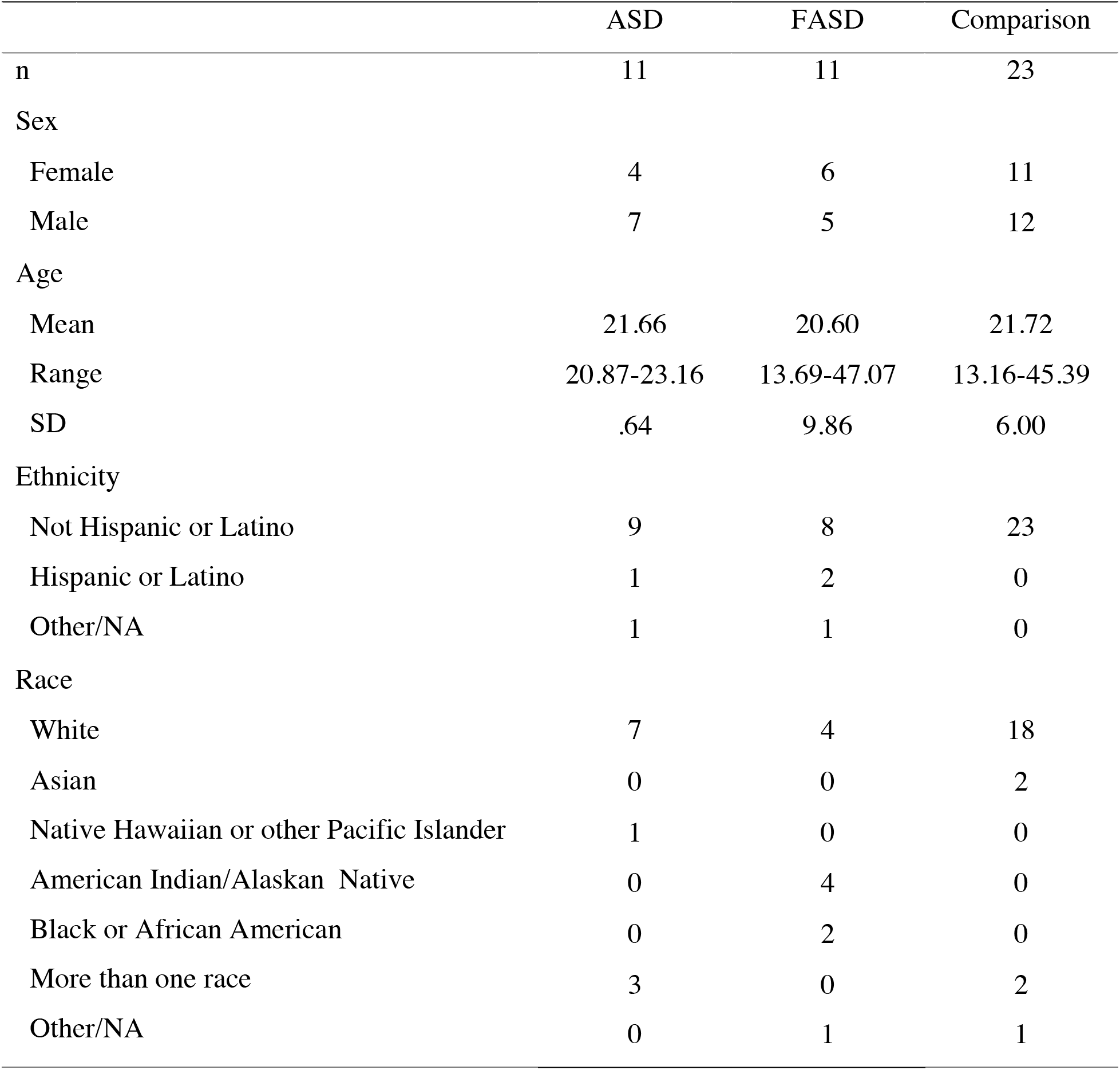
Sample Demographic Information of ASD, FASD, and comparison group

|  | ASD | FASD | Comparison |
| --- | --- | --- | --- |
| n | 11 | 11 | 23 |
| Sex |  |  |  |
| Female | 4 | 6 | 11 |
| Male | 7 | 5 | 12 |
| Age |  |  |  |
| Mean | 21.66 | 20.60 | 21.72 |
| Range | 20.87-23.16 | 13.69-47.07 | 13.16-45.39 |
| SD | .64 | 9.86 | 6.00 |
| Ethnicity |  |  |  |
| Not Hispanic or Latino | 9 | 8 | 23 |
| Hispanic or Latino | 1 | 2 | 0 |
| Other/NA | 1 | 1 | 0 |
| Race |  |  |  |
| White | 7 | 4 | 18 |
| Asian | 0 | 0 | 2 |
| Native Hawaiian or other Pacific Islander | 1 | 0 | 0 |
| American Indian/Alaskan Native | 0 | 4 | 0 |
| Black or African American | 0 | 2 | 0 |
| More than one race | 3 | 0 | 2 |
| Other/NA | 0 | 1 | 1 |

#### ASD Group

Eleven participants diagnosed with ASD were recruited from a larger longitudinal study cohort followed at the University of Washington, who were diagnosed with ASD between the ages of 3 and 4 years old. Diagnoses of ASD, according to criteria from the fourth edition (DSM-V; American Psychiatric Association, 2013), were made by a licensed clinical psychologist or supervised graduate student using: 1) the Autism Diagnostic Interview-Revised (ADI-R Lord et al., 1994), 2) the Autism Diagnostic Observation Schedule-Generic (ADOS-G; Lord et al., 1989), 3) medical and family history, 4) cognitive test scores, and 5) clinical observation and judgment (see Dawson et al., 2004; Emmons et al., 2021 for further details). These participants were evaluated again at 6, 9, and 13-15 years of age. Specific inclusion criteria for this study included normal hearing since autistic individuals have a higher incidence of hearing loss (Rosenhall et al., 1999; Szymanski et al., 2012), which could also affect language abilities. Forty-six participants from the original cohort were re-contacted and invited to participate in this current study. The remaining 26 were not contacted because they were already recruited for another study. Of the 46 participants contacted, 4 had moved out of state, 2 were not interested in participating, 1 could not be scheduled, 25 did not respond to phone calls or emails, and 2 did not meet our eligibility criteria of being able to speak in 3-word phrases. Eleven ASD participants from the original cohort were enrolled in this study and tested in their 20s (see Table 1 for sample demographic information)

#### FASD Group

Eleven participants diagnosed with FASD were recruited from Washington State Fetal Alcohol Syndrome Diagnostic & Prevention Network (FAS DPN). All participants were diagnosed using the FASD 4-Digit Diagnostic Code, which is an interdisciplinary approach to diagnose the full spectrum of outcomes guided by the magnitude of expression of the four features of FAS: growth deficiency, FAS facial phenotype, CNS structural/functional abnormalities, and prenatal alcohol exposure (Astley, 2004; see McLaughlin et al., 2019 for more details). Ten of eleven FASD participants were adolescent/young adults who were teenagers or in their early 20s; however, one participant in their 40s was also included in the study.

#### Comparison Group

The comparison group consisted of 23 neurotypical participants. For each participant in the ASD and FASD group, one age- and sex-matched participant was recruited as a pair-matched comparison participant. Age matching in comparison participants was ± 1 year with one exception. The participant in the FASD group in their 40s was matched to a comparison subject within 2 years. Participants were also matched on sex to reflect the higher incidence of males with a diagnosis of ASD. Besides the pair matched comparison subjects, there was also one additional neurotypical participants tested in their 20s. Comparison group participants all reported no history of cognitive, developmental, or other health concerns. Two participants in the comparison group had a diagnosis of ADHD but were included because there were individuals with ADHD in both ASD and FASD groups.

### Procedure

The following measures were obtained over the course of several visits and as part of a larger study that included additional neurophysiological and behavioral measures. Written informed consent was obtained from all participants or their Legally Authorized Representative for participants under 16 years of age. All methods were performed in accordance with protocols reviewed and approved by the Institutional Review Board at the University of Washington where the research was conducted. Participants were provided with monetary compensation for their time.

#### ADOS-2

The ADOS-2 (Lord et al., 2012), a measure of autism symptom severity, was administered to all participants at the time of testing to confirm ASD status in all participant groups. All participants in the ASD group received an ADOS classification of autism. No participants in the comparison or FASD group received an ADOS autism classification

#### WASI-II

The Wechsler Abbreviated Scale of Intelligence - Second Edition (WASI-II; Wechsler, 2011), a measure of intellectual ability, was administered to all participants. Four subtests, Block Design, Vocabulary, Matrix Reasoning, and Similarities were administered to compute the WASI-II Full Scale Intelligence Quotient (FSIQ-4) for each participant and provide an overall estimate of intellectual functioning. The Verbal Composite Index (VCI) score was computed from the Vocabulary and Similarities subtests and the Perceptual Reasoning Index (PRI) scores were computed from the Block Design and Matrix Reasoning subtests.

#### Auditory Screening

To ensure clinically normal hearing thresholds in all participants, each participant was required to pass an audiometric screen (≤ 20 dB hearing level at octave frequencies between 250 and 8000 Hz), a distortion product otoacoustic emission (DPOAE) screen, and an auditory brainstem response (ABR) screen.

#### Elicitation of Narratives

The 24-page wordless book, *Frog, Where Are You?* (Mayer, 1969), was used to elicit narrative language samples. This story has been used in previous research with both children and adults (Colle et al., 2008; Thorne et al., 2007) and does not require any prior descriptions or specific prompts from the examiner. This wordless picture book requires the narrator to connect and organize different actions from the protagonists and events into a coherent story, providing a measure of temporal expressions. Because the story does not provide verbal cues, the participants have the freedom to explore the characters’ inner states by taking the perspective of the characters and communicating them to the listener. The story is about a young boy and his dog in search of his lost pet frog. The protagonists’ search involves numerous events and encounters with secondary characters. Throughout the story, the two characters engage with these secondary characters in developing the story to finally find the lost pet frog.

Each participant was assessed individually, and each narration was recorded for transcription and analysis. Participants were presented with two envelopes containing copies of the wordless book *Frog Where Are You*? (Mayer, 1969). The examiner instructed the participants to review the pictures in the book to become more familiar with the story and then they were asked to use the pictures as a visual prompt to tell the best story that they could. After the participant looked through the book, the examiner confirmed that they were ready and had them turn back to the first page of the book to start recording the story. The examiner was intentionally positioned to be blocked from the view of the book. The participant went through each page of the book and told the story without any provided prompt. The examiner did not intervene during the narration. However, when necessary, the examiner was permitted to provide a verbal prompt to help the participant get started (e.g., “I will help you get started, once upon a time there was …”) or encourage the participant throughout the storytelling (e.g., “What happened next?”). This procedure ensured that the participants did not memorize the story and emphasized the storytellers’ ability to anticipate the listener’s informational needs. After the participants finished telling the story, the examiner stopped the recording and praised the participants for their storytelling skills.

#### Transcription of narratives

Narratives were transcribed orthographically and formatted *Systemic Analysis of Language Transcript* (SALT) conventions. Transcripts were segmented into *Communication units* (C-units). A C-unit is an independent clause with its modifiers, which includes one main clause with all subordinate clauses attached to it (*Salt Resources*, n.d.). As subordinate clauses depend on the main clause to make sense, they cannot be separated from the main clause, whereas the main clause can stand alone and can be segmented into C-unit. Mazes, such as filled pauses, repetitions, revisions, along with unintelligible utterances, story endings, and irrelevant comments were marked and included in the transcription. Details of the transcription can be found in Miller et al. (2008).

#### Coding of narratives

The transcribed narratives were entered into the SALT program (Miller et al., 2008), which is a software that aids the process of analyzing words, morphemes, utterances, and discourse, and were used to code and analyze structural language properties. Code from the SALT program was used to calculate the mean length of utterance, total number of utterances, total number of different word roots, total number of words, rate of speech and NSS total score. Additional code was created to measure the use of mental states in reference to temporal relations.

##### Temporal Relations

The total number of temporal expression terms, excluding mazes, were counted to test if people with ASD and FASD could organize events in sequential order in relation to the temporal organization. Temporal relations included temporal adverbs and conjunctions such as “last night”, “yesterday”, “now”, and “meanwhile”.

##### Mental States

The total number of mental state terms, excluding mazes, were counted, which included the emotional state of a character in the story (e.g., “The boy is *angry* at the dog”) and any reference to a mental state, including emphasizing with the character to explore their inner thoughts, beliefs, intentions (e.g., “The boy *thought* the frog ran away”).

##### Narrative Scoring Scheme (NSS)

Each of the narrative stories was then coded using the NSS as described in the *SALT Training Website* (n.d.). NSS assessed the individual’s ability to produce a structurally sound and coherent narrative. The scoring guideline included many features of Story Grammar, cohesiveness, connection of story events, consideration for characters’ thoughts and behaviors, and referencing. Each story was graded on a rubric of 7 categories with a grading scale of 0-5: introduction, character development, mental states, referencing, conflict/resolution, cohesion, and conclusion. Categories that could not be scored received a score of zero or N/A. Scores of zero were given for the target speaker errors (e.g., not following the protocols or telling the wrong story) and a mark of non-applicable (N/A) was given for the examiner errors (e.g., interference with the speaker’s story or recording issues). For each section, a score of 1 indicated minimal/immature performance, a score of 3 indicated emerging skills, and the highest score of 5 indicated proficient characteristics. Scores of 2 and 4 were undefined and given based on the examiner’s judgment with justification. The scores for each section were then combined to give a total narrative score with the highest possible score of 35. For more details on scoring and guidance using the NSS, see the SALT Training Website (n.d.).

#### Interrater agreement

To ensure the reliability of the data, a systematic process of transcription and scoring was designed to prepare all narratives from recorded sourced files by two researchers with 100% of the narratives being conducted by a secondary researcher who was not informed of the group identity or diagnosis of the narrator. Inter-rater reliability was calculated for all measures with 93% agreement on words and 89% agreement on C-units in terms of transcription and 96% agreement on each category of the NSS total score. Percent agreements were calculated based on the number of differences in terms of the numbers of words, scores between the primary and secondary coder. The second independent researcher’s final score for each category of NSS was used to eliminate author’s biases.

## Results

First, narrative performance of the ASD and FASD groups were compared to their matched comparison groups. One-way analysis of variance (ANOVA) was used to test for statistical differences between the means of all the groups. No significant differences were detected between groups for any measure; thus, the comparison groups were combined as a single group for all subsequent analyses.

### Local Measures

We investigated the difference between participant groups in terms of mental states and temporal relation words. Table 2 presented the means and standard deviations for two measures of local abilities. One-way ANOVA revealed no statistically significant group differences in the number of mental state words produced (Figure 1B; one-way ANOVA, *F*(2,42) = .54, *p* = .59, *η*^*2*^ = .02) or temporal relation words produced (Figure 1A; one-way ANOVA, *F*(2,42) = 1.85, *p* = .17, *η*^*2*^ = .08).

**Table 2.** Local Measures Raw Scores: Means, Standard Deviations, and One-way ANOVA Comparisons between ASD, FASD, and NT Comparison Groups.

| Local Measures | ASD | | FASD | | Comparison | | $F(2,42)$ | $\eta^2$ |
| --- | --- | --- | --- | --- | --- | --- | --- | --- |
| | $M$ | $SD$ | $M$ | $SD$ | $M$ | $SD$ | | |
| Temporal Relation Words | 11.45 | 7.29 | 8.09 | 3.33 | 8.13 | 4.30 | 1.85 | .08 |
| Mental State Words | 6.82 | 2.87 | 5.45 | 3.46 | 5.83 | 3.46 | .53 | .02 |

**Figure 1.**
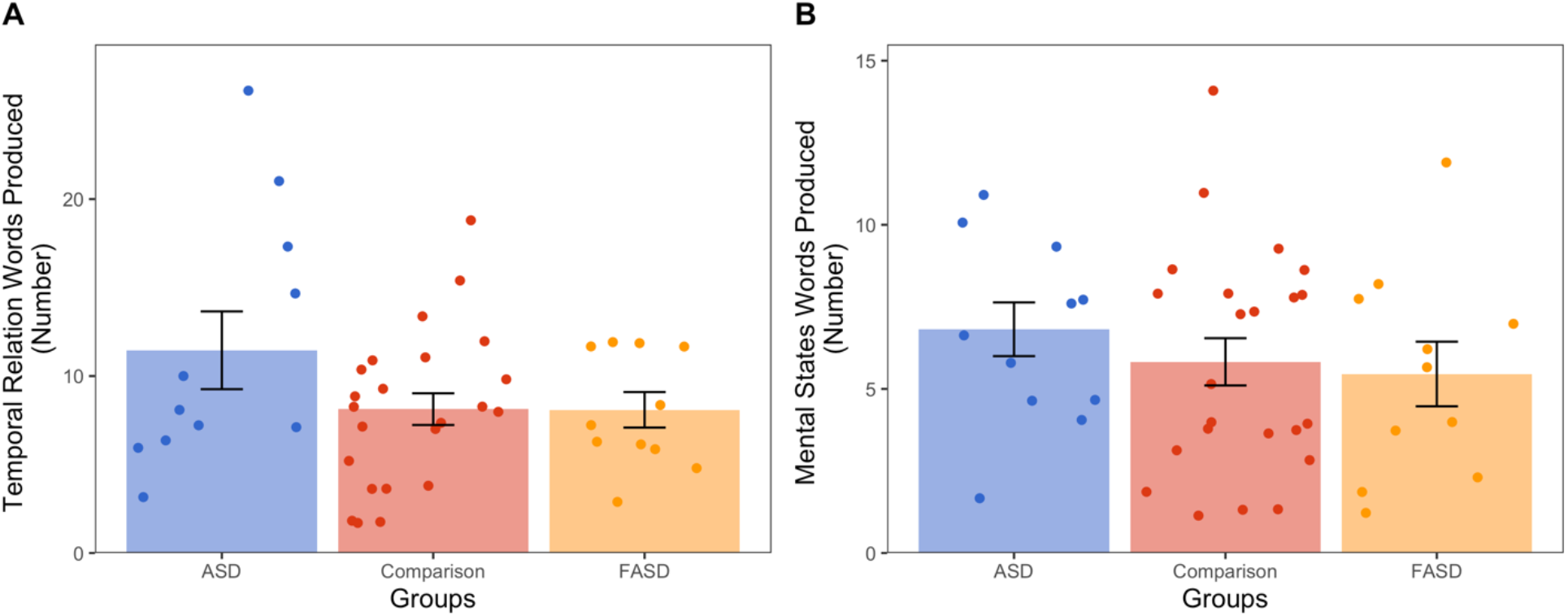
Local measures: temporal relation words (A) and mental state words (B), as a function of the participant group. Mean ± SE is shown with bars. Individual data points are shown with solid circles. No group differences were observed for the number of temporal relation and mental state words produced.

#### Relationships between local measures and global measures

Pearson correlation coefficient (Pearson’s *r*) measured the strength of the linear relationship between temporal relation words produced and other global measures. This revealed a positive correlation between the numbers of total words (including mazes) and the number of temporal relation words (Pearson’s *r, r*(43) = .60, *p* < .001). The longer the story produced, the greater number of temporal relation words observed.

### Global Measures

We investigated the difference between participant groups in terms of global measures. Table 3 presents the means and standard deviations for five measures of global abilities. One-way ANOVA comparing each global measure in ASD, FASD, and comparison group revealed no significant group differences on any of the measures besides the total number of words including mazes and rate of speech. Additionally, Pearson’s *r* was conducted to measure the strength of the linear relationship between global variables and indicated a strong correlation between the number of total words produced and the NSS total score as well as the number of different words produced and NSS total score.

**Table 3.** Global Measures Raw Scores: Means, Standard Deviations, and One-way ANOVA Comparisons between ASD, FASD, and NT Comparison Groups.

| Global Measures | ASD | | FASD | | Comparison | | $F(2,42)$ | $\eta^2$ |
| --- | --- | --- | --- | --- | --- | --- | --- | --- |
| | $M$ | $SD$ | $M$ | $SD$ | $M$ | $SD$ | | |
| Total Words with Mazes | 394.09 | 133.93 | 314.09 | 124.15 | 283.91 | 97.31 | 3.51* | .14 |
| Words/Minute | 127.65 | 17.67 | 115.19 | 28.24 | 159.04 | 20.43 | 17.37*** | .45 |
| Total Different Words | 118.36 | 31.19 | 106.55 | 27.03 | 104.57 | 26.42 | .95 | .04 |
| Mean Length of Utterances | 11.00 | 2.80 | 9.74 | 1.72 | 10.40 | 1.89 | .98 | .04 |
| Total Utterances | 32.00 | 10.55 | 30.82 | 14.21 | 25.87 | 7.09 | 1.75 | .08 |
| NSS Total Score | 24.91 | 5.30 | 22.27 | 6.42 | 22.74 | 4.60 | .84 | .04 |

\* $p < .05$
\*\*\* $p < .001$

#### Total Number of Words (including Mazes)

The number of total words produced including mazes showed that a longer narrative story was observed for ASD group compared to FASD and comparison group (see figure 2A). One-way ANOVA revealed a significant group difference in total numbers of words, including mazes (one-way ANOVA, *F*(2,42) = 3.51, *p* = .04, *n*^*2*^ = .14). Post hoc comparisons were conducted using independent sample t-Tests with Bonferroni correction, which revealed a significantly higher number of words produced in the ASD group (*M* = 394.09, *SD* = 133.93) compared to the comparison group (*M* = 283.91, *SD* = 97.31). However, no significant group difference were observed between the ASD and FASD groups or between the FASD and comparison groups.

**Figure 2.**
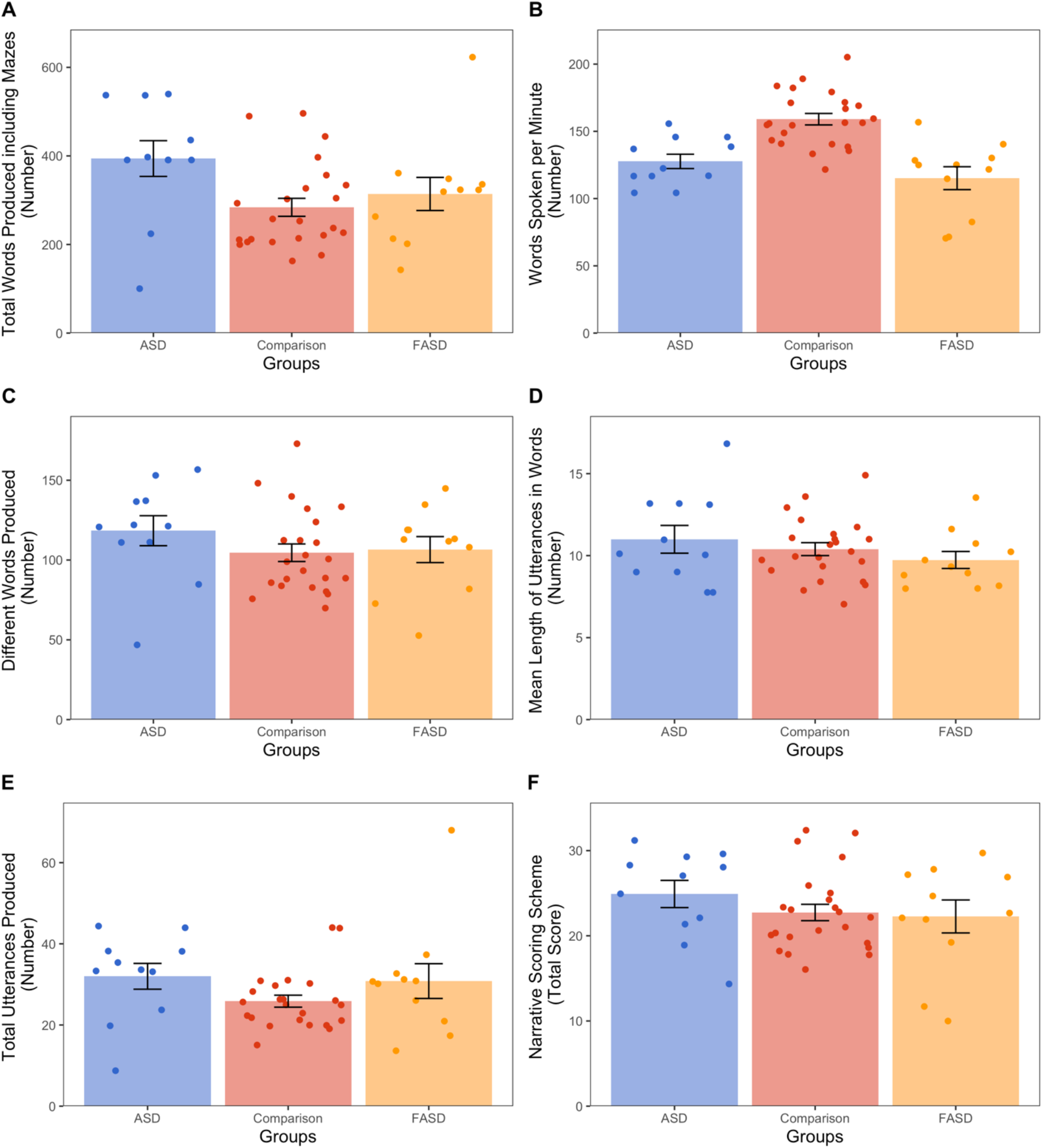
Global measures: total number of words produced (A), number of words spoken per minute (B), number of different words produced (C), mean length of utterances in words (D), total number of utterances produced (E), and the NSS total score (F) as a function of the participant group. Mean ± SE is shown with bars. Individual data points are shown with solid circles. Group differences were observed only for total number of words produced (including mazes) and words spoken per minute.

#### Rate of speech

The comparison group produced speech at a faster rate, which is the number of words spoken per minute, compared to ASD and FASD groups (see figure 2B). One-way ANOVA revealed a significant difference across groups in the number of in words spoken per minute (one-way ANOVA, *F*(2, 42) = 17.37, *p <* .01, *n*^*2*^ = .45). Post hoc comparisons using independent sample t-tests with Bonferroni correction, revealed a lower rate of speech in the ASD (*M =* 127.65, *SD* = 17.67) and FASD (*M* = 115.19, *SD* = 28.24) groups compared to the comparison group (*M* = 159.04, *SD* = 20.43). However, no significant difference was observed between the ASD and FASD group.

#### Relationships within global measures

We investigated the relationships between global variables, which were presented in Table 4. A strong correlation between the number of total words produced and the NSS total score (Pearson’s *r, r*(43) = .74, *p* < .001) was found, indicating that the more words produced, the higher the NSS total score. A strong correlation between the number of total different words produced and the NSS total score (Pearson’s r, *r*(43) = .89, *p* < .001) was also found, indicating that the more different words produced, the higher the NSS total score.

**Table 4.** Correlations for Global Measures.

| Global Measures | 1 | 2 | 3 | 4 | 5 | 6 |
| --- | --- | --- | --- | --- | --- | --- |
| 1. Total Words with Mazes | — |  |  |  |  |  |
| 2. Words/Minute | .04 | — |  |  |  |  |
| 3. Total Different Words | .89*** | .14 | — |  |  |  |
| 4. Mean Length of Utterances | .40** | .36* | .60*** | — |  |  |
| 6. Total Utterances | .81*** | -.12 | .63*** | -.16 | — |  |
| 7. NSS Total Score | .74*** | .16 | .84*** | .51*** | .52*** | — |

\* $p < .05$
\*\* $p < .01$
\*\*\* $p < .001$

## Discussion

In this study, we elicited narratives using a wordless picture book to investigate narrative discourse abilities in a sample of adolescents and adults with ASD, FASD, and an age- and sex-matched comparison group. We evaluated both local and global language features of the narratives produced. Important differences were observed in the number of total words produced and rate of speech. ASD group produced significantly more total words compared to the comparison group and the ASD and FASD groups had a lower rate of speech compared to the comparison group. No significant group differences were found in local measures such as the use of temporal relation and mental state terms. Our findings also suggested that many aspects of global narrative production, including total number of different words, total number of utterances, mean length utterance, and the NSS total score in the ASD and FASD groups were comparable to the comparison group and to each other.

The control of local and global language features contributes to successful social communication. Moreover, narrative storytelling also requires the speaker to consider a listener’s perspective to construct a cohesive narrative as they organize the story. Our results provided answers to the broad question: are there differences in the local and global structure of narratives produced by participants with ASD, FASD and the comparison group? Each of these findings is discussed in more detail below.

### Local Measures

#### Temporal Relations

There was no significant group difference observed in the number of temporal relation words produced by the ASD, FASD, and comparison groups. This finding is in contrast with Colle et al. (2008) that did find a difference between their ASD and comparison groups. In our participant sample, the production of temporal relation words in individuals with both ASD and FASD were comparable to the comparison group, which is when storytellers were expected to consider the story from the perspective of the listeners and use the relation between events to make sense of the storyline. Additionally, a positive correlation between the numbers of total words (including mazes) and the number of temporal relation words was observed, which indicates that the number of temporal words produced is proportional to the length of the story itself but may not necessarily tell us about how participants maintain and use temporal references. The ability to maintain and use different types of temporal references in a narrative in an efficient way should be further assessed in future studies.

#### Mental States

In line with the findings from Colle et al. (2008) and Tager-Flusberg and Sullivan (1995), the results did not show any significant differences between the ASD, FASD, and comparison groups in the production of mental state words. This, however, was not what we predicted. These findings are consistent with some of the results from narrative abilities of autistic individuals.

For example, Colle et al. (2008) found that adults with ASD did not show any significant difference in the use of mental state expressions. Similarly, Tager-Frusberg and Sullivan (1995) showed that children with autism produced a similar quantity of emotional and mental state words. However, they noted that the children with ASD were more likely to use expressions with a limited understanding of intentions and internal states of the character or taking into consideration the point of view of the listener. In short, these children were able to label emotions but did not fully understand the role of the mental state expressions that they used. This finding suggests that although autistic individuals may produce the same number of mental state words as the comparison group, they may show deficits in comprehension that are not captured by the measures employed in this study.

### Global Measures

The results showed significant group difference between ASD, FASD, and comparison subjects on the length of narrative (total words including mazes) produced and rate of speech. Post hoc analyses revealed a greater number of total words produced by the ASD group compared to the comparison group. However, no significant group differences were found between the ASD and FASD or between the FASD and comparison group.

Additionally, a lower rate of speech was observed in both ASD and FASD groups compared to comparison subjects, but not but no differences between the ASD and FASD groups were observed. These findings are inconsistent with past studies that found comparable length of narrative between ASD and comparison groups (Banney et al., 2015; Colle et al., 2008; King et al., 2014; Loveland et al., 1990; Tager-Flusberg & Sullivan, 1995).

On other aspects of the global features, the results showed that the ASD and FASD groups were comparable to each other as well as the comparison group, including the mean length of utterances, total number of utterances, total number of different words produced, and the NSS total score. This result is consistent with Reindal et al. (2021) and Colle at al. (2008)’s finding that children with ASD had comparable structural language skills to children without ASD such as choice of vocabulary. However, since pragmatic language impairments were found to be most salient in children with ASD compared to children without ASD and is an essential component of social communication, pragmatic language skill should be examined in relation to the structural components of narrative production.

One strength of the study is the multi-group examination approach, which enabled us to compare the narrative language skills in individuals with ASD and FASD. Such examination increases the generalizability of findings to the broader population of adults evaluated for ASD and FASD since the literature on narrative language abilities in FASD is still limited.

One limitation of the study is the small sample size (*n* = 11 ASD; *n* = 11 FASD; *n* = 23 comparison). For this reason, only group level analyses were conducted. While our findings suggest strong narrative abilities for FASD and ASD groups as a whole, there is also significant variability across individuals. Such results suggest that language abilities need to be assessed at the individual level. Given the widely distributed performance in both local and global levels of narrative language production, future studies should also assess their subjects based on their groups’ distinctive characteristics such as attention deficit and compulsive nature in individuals with FASD (Nanson & Hiscock, 1990) and consider their additional variables that influence narrative production, such as motivation, distractibility, decision making of individual subjects. Additionally, since only one storytelling task was obtained in this study, the assessment does not reflect how each individual would perform across different settings such as daily social communication versus structured language assessment. Although the sample did not show a significant difference on this particular task, they may show deficits on other higher language measures. Furthermore, although the findings of the study may be useful in identifying the narrative language abilities between individuals with ASD and FASD with this form of narrative discourse, they may not generalize to other individuals with ASD and FASD or other types of narratives. Additional tasks and analyses involving narrative storytelling would provide useful comparisons between these two groups. Lastly, when we integrate the ability to use context to support other structural components of language (e.g., providing temporal relations terms to make a smoother transition between parts of a story; using non-linguistic signals to communicate (Barokova & Tager-Flusberg, 2020), we perhaps use more than just spoken language to deliver our stories. As such, future efforts in understanding narrative discourse in adults with FASD and ASD should be extended to all components of language and especially, expressive, or pragmatic language skills.

Given the limited number of past studies on the narrative skills of adolescent/young adults with FASD and their shared characteristics with autism (Stevens et al., 2013), this present investigation of narrative storytelling will contribute to what we know about the language abilities of individuals with FASD. This study suggests that many aspects of narrative production in individuals with ASD and FASD are comparable to the comparison subjects and to each other when considered at the group level. This includes both local and global features. With the rich empirical research available in autistic individuals, the results highlighting the similarities between ASD and other neurodevelopmental conditions like FASD could lead to more research that is focused on the representations of autistic characteristics in other neurological conditions (e.g., Stevens et al., 2013), and specifically its potential influence on the development of language skills. Ultimately, the results could aid in the development of clinical tools to best assess language abilities in individuals with FASD. Eventually, this understanding could also lead to the development of language support systems that further help individuals advance their language skills and gain self-confidence in the daily social communication (Nanson & Hiscock, 1990). Finally, the current assessments protocols used to diagnose language disorders should be reconsidered to include narrative assessment to avoid underestimating language abilities. Lastly, this study only included adolescent/young adults, future research should compare the development of language skills from childhood to adulthood in individuals with ASD and FASD.

## Summary

Our results suggest that many aspects of narrative production in individuals with ASD and FASD are comparable to the comparison subjects but that important differences are observed in the total number of words produced and rate of speech. Specifically, the ASD and FASD groups both spoke at a significantly lower rate of speech compared to the comparison group and individuals in the ASD group produced significantly more words compared to the comparison group. Given that significant individual variability within groups was observed, the findings also suggest that language abilities should be assessed at an individual level and future research should consider additional variables that influence narrative production such as motivation, distractibility, or decision making of individual subjects. Taken together, this current study contributes one aspect in understanding the multifaceted components of language abilities, and provides broad theoretical and real-world implications for the development of clinical assessment and educational support for individuals with ASD and FASD.

## Data Availability

All data produced in the present study are available upon reasonable request to the authors

## Acknowledgements

We thank the subjects and their families for their participation in this research.

## Data Availability

All data and analysis code are available from Bonnie K. Lau at upon request.

